# Health Care Provider Compliance with the HIPAA Right of Individual Access: a Scorecard and Survey (Revised)

**DOI:** 10.1101/19004291

**Authors:** Deven McGraw, Nasha Fitter, Lisa Belliveau Taylor

**Affiliations:** Ciitizen Corporation

## Abstract

**Background:** Historically, patients have had difficulty obtaining copies of their medical records, notwithstanding the legal right to do so. In 2018, a study of 83 top hospitals found discrepancies between those hospitals’ published information and telephone survey responses regarding their processes for release of records to patients, indicating noncompliance with the HIPAA right of individual access.

**Objective:** Assess state of compliance with the HIPAA right of access across a broader range of health care providers and in the context of real records requests from patients.

**Methods:** Evaluate the degree of compliance with the HIPAA right of access 1) through telephone surveys of health care institutions regarding release of records to patients and 2) by scoring the responses of a total of 210 health care providers to actual patient record requests against the HIPAA right of access requirements. (51 of those providers were part of an initial cohort of 51 scored for an earlier version of this paper.)

**Results:** Based on the scores of responses of 210 health care providers to record requests and the responses of nearly 3000 healthcare institutions to telephone surveys, more than 50% of health care providers are out of compliance with the HIPAA right of access. The most common failure was refusal to send records to patient or patient’s designee in the form and format requested by the patient, with 86% of noncompliance due to this factor. The number of phone calls required to obtain records in compliance with HIPAA, and the lack of consistency in provider responses to actual requests, makes the records retrieval process a challenging one for patients.

**Conclusions:** Recent federal proposals prioritize patient access to medical records through certified electronic health record (EHR) technology, but access by patients to their complete clinical records via EHRs is years away. In the meantime, health care providers need to focus more attention on compliance with the HIPAA right of access, including better training of staff on HIPAA requirements. Greater enforcement of the law will help motivate providers to prioritize this issue.

## Introduction

In October 2018, researchers affiliated with Yale University published a study in JAMA Open Network evaluating the processes at 83 top hospitals for responding to patient requests for their medical records under the Health Insurance Portability and Accountability Act (HIPAA) Privacy Rule.[1] The researchers called institutions and inquired about their processes for getting records to patients and compared them to published information about those processes. The study found discrepancies between the records release processes described in the request forms and the description of the process given by institution staff by phone, which indicated noncompliance with HIPAA (as well as applicable state patient record laws).

The study resonated with our own experiences helping the beta users of Ciitizen obtain their medical records. Ciitizen is a new consumer company developing a personal health record platform to enable patients, beginning with cancer patients, to obtain all of their health information under their HIPAA right of access, allowing them to then share that data to seek second opinions, determine eligibility for clinical trials, and donate data for research. As we began to help the initial beta users of our platform to obtain their medical records using their HIPAA right of access, we recognized widespread noncompliance among health care providers with the HIPAA right of access.

We submitted HIPAA medical requests for records to 210 health care providers. These were legitimate access requests, on behalf of users who had consented to opening Ciitizen accounts and to having us help them access their health information for the purpose of populating those accounts. We then scored those experiences in comparison to both what is required by the HIPAA right of access, and whether any providers went above and beyond to get patients their records more promptly via a seamless process. As expressed in more detail below, 51% of these providers are either not compliant with the HIPAA right of access or were compliant only after two or more phone calls to privacy officials or health information management (HIM) supervisors to educate them about HIPAA’s requirements. Among those providers who were compliant with HIPAA’s requirements, we found 20% to be going above and beyond what the law requires.

In preparation for submitting those requests, and using a process similar to that used by the Yale researchers, we surveyed, by telephone, thousands of hospitals regarding their processes for releasing medical records pursuant to a patient request. The survey results for nearly 3000 of those hospitals indicated that as many as 56% of providers could be out of compliance with HIPAA.

## Methodology

### The Scorecard: Process

The beta users of the Ciitizen platform were individuals either known to or related to Ciitizen staff, were referred by individuals or patient advocacy organizations known to Ciitizen staff, or they signed up on a waitlist maintained on the Ciitizen website. Between February 10, 2019 through September 30, 2019, we submitted written medical records requests, using a HIPAA-compliant form developed by Ciitizen, to providers specified by each user, covering a specific timeframe, requesting all medical records, including images, generated within that timeframe. The records requests were signed by the user (and accompanied by a photo of the user’s driver’s license, for purposes of proving identity) and submitted by email or by fax, depending on the process acceptable to the provider. The request also indicated whether the patient further consented to having certain types of sensitive data (for example, genetic information, reproductive health information, HIV/AIDS, and substance abuse treatment information) sent to Ciitizen, because many providers believe that state or other federal laws require this additional acknowledgement, even for sharing with patients. [2] The request specified that the information be sent directly to Ciitizen either through upload into a secure portal or by email and, if by e-mail, expressly acknowledged and accepted the security risks of receiving information unsecurely. (This is required by HIPAA for individuals seeking that their data be sent by unsecure email. [3]) The request also asked for an estimate of any fees associated with completing the request.[4]

Ciitizen’s outreach process to medical records offices involved multiple phone calls to:

- Confirm the request for records was received;
- Follow up, answering questions explaining HIPAA requirements; and
- Escalate to supervisors and/or privacy officers where there were indications HIPAA might not be followed.

Staff took careful notes of what occurred during the process, from submission of the request to fulfillment.

### The Scorecard: Scoring

The providers to whom the record requests were sent were scored based on one to five stars. The first four stars measure their compliance with core requirements of the HIPAA Privacy Rule right of access, as articulated in the HIPAA privacy regulations and guidance issued by the federal Department of Health and Human Services (HHS) Office for Civil Rights (OCR), the office with authority to promulgate policy under and enforce HIPAA’s privacy mandates.[5] [6] (Of note: although there are a number of state laws that set a higher bar for patient access to records, we evaluated only compliance with the HIPAA Privacy Rule.) Specifically:

- <u>Provider accepts requests by email or fax</u>: Providers may not create a barrier to access by requiring patients to submit requests in person or by mail.[7]
- <u>Records were sent in the format requested to the patient’s designated recipient:</u> The provider sends the records in the format the patient requests, which is in digital form by email (or upload to portal) for text), and sends it to the third party designated by the patient.[8]
- <u>Records were sent within 30 days:</u> The provider responds to the request within 30 days of receipt (or, if within 30 days they provided a written statement of reasons for the delay and the date by which the records would be provided, the records were received within 60 days of receipt of the request).[9]
- <u>No unreasonable fees charged for the request:</u> Providers may only charge reasonable, cost-based (i.e., minimal) fees to cover labor costs of copying and supplies.[10]

Providers received one to two additional stars for having seamless processes and for going above and beyond what HIPAA requires to get patients their records. More details on the scoring methodology can be found in Box 1. Of note: we did not score the ease and completeness of obtaining records through electronic medical record portals or application programming interfaces because the amount of records the patient has a right to under HIPAA is not yet available through these mechanisms.

Most providers were scored only on a single request; for providers receiving more than one request, we scored only the most recent request, as an indication of either improvement or, ideally, consistency in responding to patient requests.

#### Box 1

**Scorecard Scoring**

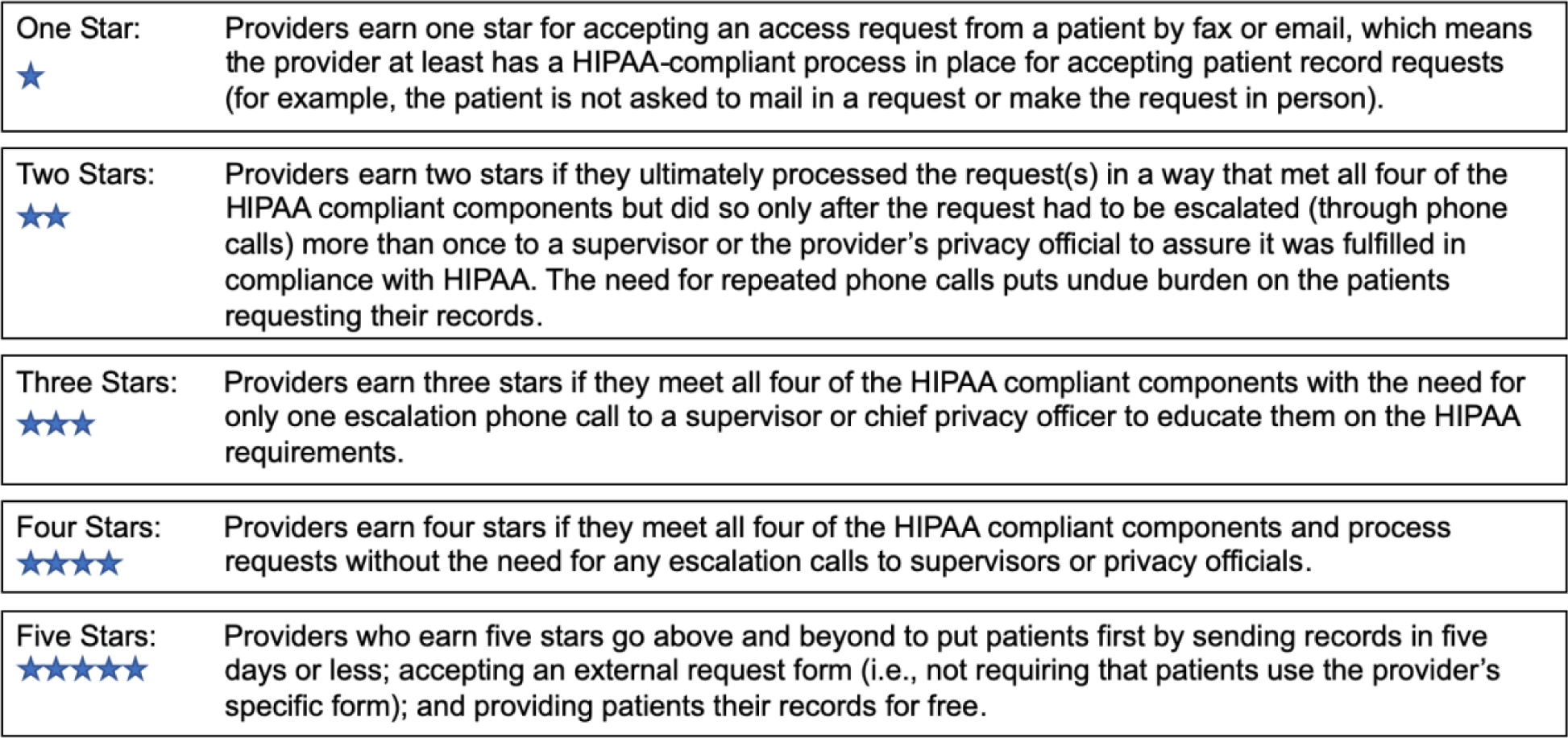

In August 2019, an initial version of this paper reported scores for an early cohort of 51 providers; this revised paper reports on scores for a total of 210 providers, 51 of whom were also part of the initial cohort. The scores for each provider can be found at www.ciitizen.com/scorecard.

### The Survey: Process

In preparation for sending out requests for access on behalf of our users, we searched for a directory of all hospitals and health systems in the U.S. with information about their patient record access processes. We started with hospitals and health systems because they are repositories for a large amount of medical information needed by a cancer patient. No such database currently exists, so we set out to create it. We first attempted to use the Medicare National Provider Identifier (NPI) Database, but found it riddled with duplicates and missing information needed for our purposes. We garnered names of hospitals and health systems by doing Internet searches using phrases such as “top hospitals,” “largest health systems in the US,” “largest hospital systems in America” “top/largest for-profit hospitals,” “top / largest non-profit health systems”,”top medical centers” and “top cancer centers.” Between August 2018 and May 2019, we made phone calls to thousands of hospitals and health systems and had reportable data on approximately 3000. (A large number were difficult to reach by phone (no answer and no return calls to voicemails left on machines), and for others the respondents did not know the answers to the questions, or the responses were too confusing to report or were not reliably recorded.)

Because we were just gathering information to build a database and were not initially setting out to do a systematic investigation of institution responses, the selection is not representative, nor does it constitute a random sample. However, for each institution we used the same script to gather information, with the questions asked matching the HIPAA right of access requirements (see Box 2 for our process and a sample script together with the questions posed to each institution). We realized after building the database that the information constituted an informal survey of hospital and health system patient access request processes and decided there was value to the public in publishing it.

#### Box 2

**Survey Process**

1. Call the main switchboard, or, via website information find a direct phone line to the medical records or health information management department. Record number that reaches a live person.
2. Ask “If a patient is out of state and needs copies of their medical records, will you accept a fax or emailed authorization form that includes a copy of their ID and signature?” Record the information as a Y or N; we also recorded the fax and/or email address as appropriate. (We asked the question as an out-of-state patient to assure that we received a response appropriate for remote (not in-person) access.)
3. Ask “If the patient is requesting their medical records be sent electronically (such as by email), are you able to send them their records in that way?” Record the response as Y or N; record N if they refuse to send records to the patient by any means.
4. Ask “Do you charge the patient for their medical records?” If they do charge, ask how much or how they arrive at the charge and record the details.
5. Ask whether radiology imaging can also be requested through the Medical Records Department or if the request needs to be separately made to the Radiology Department/Film Library. For those institutions that indicated they release images only through their Radiology Departments/Film Libraries, call the Radiology Department/Film Library and ask:
  a. If a patient is out of state and needs copies of their actual images, will they accept a fax or emailed request form that includes a copy of their ID and signature?
  b. If they would mail images to patients on a CD (images are too large to send by email).
  c. If they charge patients for images, and if so, how much or by what methodology do they determine the charges.

Institutions were evaluated whether they indicated on the phone that they would accept a request for access (for both records and images) by fax or by email; whether they would send records by email, or images by CD; and whether their purported fees for access were within the bounds of what HIPAA permits. All of these questions address four key aspects of patients right of access under the HIPAA Privacy Rule:

1. the right of a patient to receive records directly (versus sending records only to another health care provider) [5];
2. the right of a patient to submit a request in ways that do not cause undue delay or impose a burden [11];
3. the right of a patient to receive records in the form and format requested, including receiving electronic text records by email [3]; and
4. the right to have any fees for these records be reasonable (reasonable, cost-based fees for the labor needed to make the copy) [12].

Based on their responses, we evaluated whether their responses indicated compliance with HIPAA. Hospitals were deemed to be likely in compliance with HIPAA if their responses to all of the questions were consistent with HIPAA compliance; a noncompliant answer to any question earned the hospital a “no” (N) in the category of indicated compliance with HIPAA.

With respect to fees, the HIPAA Privacy Rule permits health care providers to charge only reasonable, cost-based fees to cover labor costs of copying and any associated supplies.[12] In guidance, OCR sets forth three options for calculating the appropriate reasonable, cost-based fee for the labor associated with making the copy: 1) calculating the actual fee for each request, 2) establishing a fee schedule, such as based on the size of the file, or 3) an easy to apply flat fee of up to $6.50 for digital copies of electronic health records. [13]] OCR guidance also makes clear that per page fees, which are often set forth in state law, are not permitted to be charged for digital copies of digital records.[14]

We evaluated the institutions’ responses on fees in the following way:

- We considered an institution to likely be charging “reasonable fees” for patient access if in their responses they stated that they:
  - did not charge patients,
  - charged a flat fee of $6.50 or less for a digital copy (including a copy on CD, even though we were not asking for records in that format), or
  - reported fees that seemed to be based on reasonable labor costs for copying (for example, by responding that the costs were $X per hour of copying).
- We considered an institution to likely be charging “unreasonable fees” if in their responses they stated that they:
  - charged per page fees (even if initial pages were free), including any fees for records retrieval, or
  - charged a flat fee higher than $6.50.
- Institutions that did not answer this question, or whose responses were too confusing to evaluate, are reported as NA (not applicable). We removed all institutions with NA from the denominator in calculating the percentage of institutions whose responses indicated compliance with this aspect of the HIPAA right of access.

Because this is based on phone inquiries, and not a response to an actual patient request, we were unable to evaluate whether records would be provided within HIPAA’s 30-day timeline or whether the records would be sent to a third party designee.[15] [16] We are also deeming these responses to be “indications” of compliance or noncompliance because they are not responses to an actual records request submitted by a patient.

The analysis section reports some overarching trends among the institutions who responded to our queries. Detailed survey results for each institution surveyed can be found at www.ciitizen.com/scorecard.

## Analysis

### The Scorecard

210 healthcare providers were scored based on how they processed an individual HIPAA right of access request for a Ciitizen user. (Figure A) 95 of those providers - 45% - received only one star because they failed to meet all of the basic components of the right of access except for having a compliant process for accepting patient requests (by e-mail or by fax). An additional 13 providers received two stars, which means they met the basic components of HIPAA but needed significant prompting about the HIPAA requirements in order to do so (two or more phone calls to supervisors or privacy officials). Consequently, 108 providers - 51% - were either out of compliance with the HIPAA right of access or met those requirements only after escalation to higher level officials at least twice (which suggests that a patient request not accompanied by learned intervention would not have been met with a compliant response). On a positive note, 40 percent of scored providers — nearly half — received either 4 or 5 stars.:

**Figure A:**
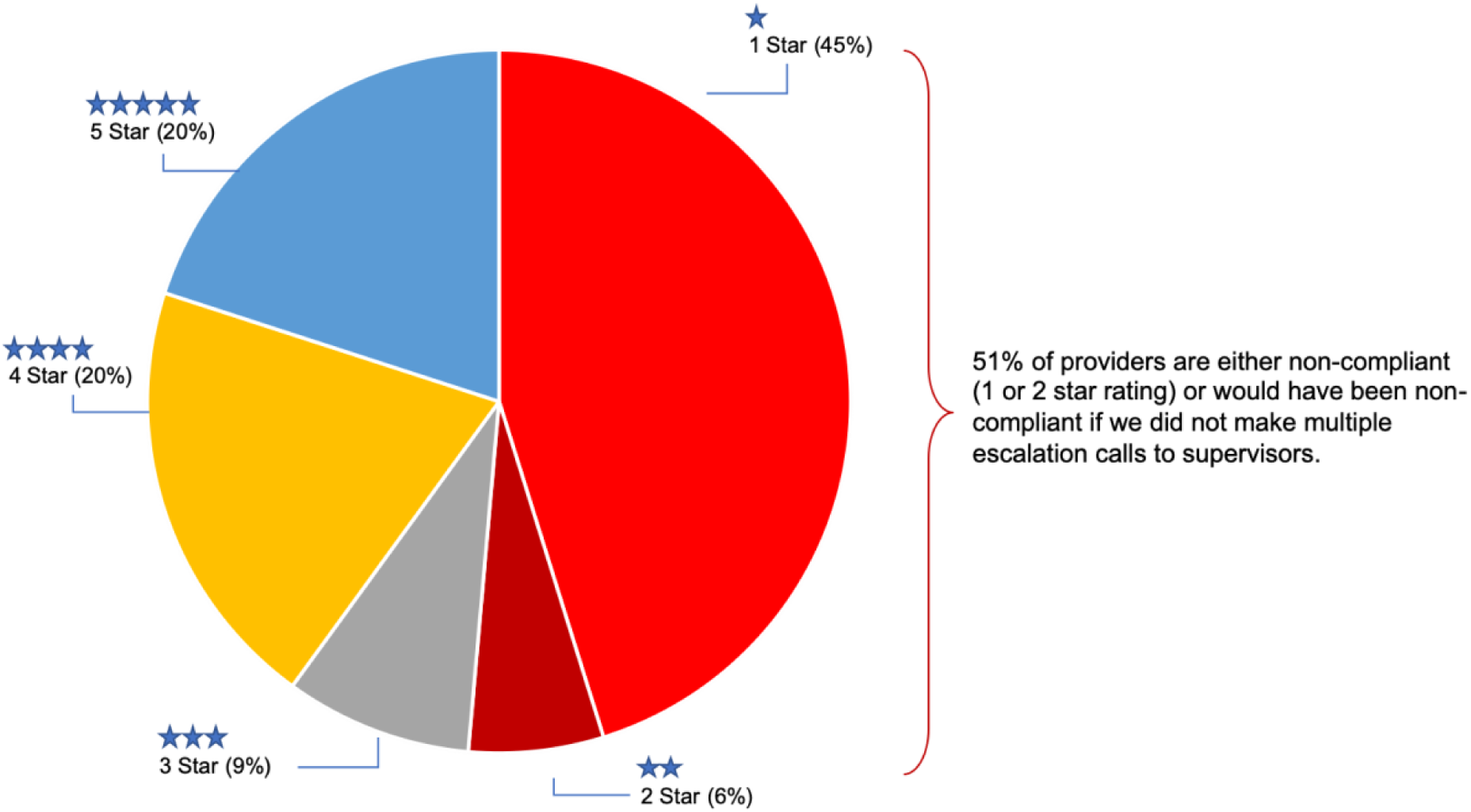
Compliance with HIPAA. (n=210 Healthcare Providers) Compliance with HIPAA based on 210 healthcare providers

### The primary reason for noncompliance is that healthcare providers do not send records in the form and format requested by the patient

Of the 95 healthcare providers receiving one star, 82 of them (86%) failed for not providing records in the electronic form and format requested by the patient (by upload to secure portal or unsecure email for text records). 17 providers (20%) were noncompliant for failing to provide records within 30 days and only two providers (2%) were noncompliant for charging unreasonable fees (one was noncompliant for failure to send to the person or entity designated by the patient). Providers and their copy services continue to send paper records, encrypted emails, faxes and CDs - even when the patient explicitly requests records be uploaded to a secure portal or sent electronically to a designee over standard email. Healthcare providers are also hesitant to send records by standard (unsecure) email, even pursuant to specific patient requests that include acknowledgement and acceptance of security risks. Figure B below shows the various ways patient records were sent.

**Figure B:**
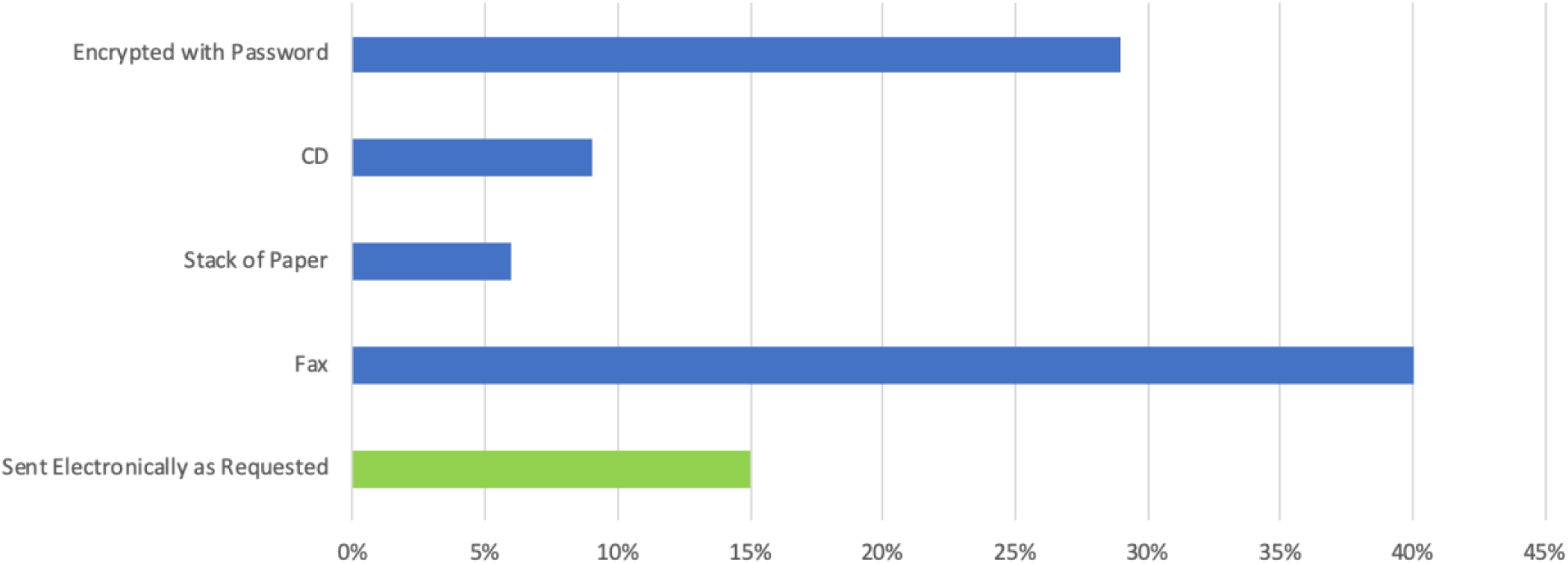
Methods of Delivery of Medical Records. (n=95 Healthcare Providers Receiving One Star) Methods of delivery of patient records requests

### Many more providers would be compliant but for disregarding patient requests re: form and format

Out of the 95 providers who were not HIPAA compliant, 71 (75%) would have been HIPAA compliant if we gave them credit for sending the information in any format other than paper (e.g., by encrypted e-mail, CD or fax). If we disregarded form and format for all but paper, 67 providers (31.9%) would have been five-star providers (the information was sent in under five days, without requiring the patient to complete a specific form, and for free); an additional 67 (31.9%) would have been four star providers (seamless, compliant process); 32 (15.2%) would have been three star providers (one escalation call), two (9.5%) would have been two star providers (two or more escalation calls), and only 24 (11.4%) would have been noncompliant.

As a company, Ciitizen can handle data in any format sent to us, as we have a fax machine, CD readers, and a scanner. It is tempting to reward providers who are getting records to patients or their designees seamlessly and promptly, regardless of the format – especially since for so many providers, the form/format requirement is the only issue that prevents them from being rated at either 4 or 5 stars. But ultimately, the scorecard reflects actual HIPAA compliance. “Form and format” can be important to patients, who often can’t accept a fax or CD or for whom encrypting data could create a barrier, because the encryption can “stick” to the data and the passwords typically will expire in 30 days or less. OCR’s guidance emphasizes that the patients can choose convenience over security in getting their records, and providers (or their vendors) who ignore this aspect of a patient’s request are placing obstacles in the path of patients exercising their HIPAA Right of Access.

### Providers are not consistent in responding to record requests

Although most providers on the scorecard were scored based on only one request, 16 providers have received multiple (two or more) requests, and their scorecard rating is based only on their last request as a measure of consistency. Of the 16, only 4 (25%) showed improvement over time – the remaining 12 (75%) saw their scores decline, had high variability (from high performance to low performance, or with one instance of high performance sandwiched between lower scores), or had consistently poor scores (1 or two stars). Because OCR can launch an investigation based on one patient’s request out of compliance with the law, it is critical that providers be consistent in handling patient requests in compliance with the law.

### The Survey

We conducted a phone survey of thousands of health care institutions to assess likelihood of compliance if patient requests were made to their offices (see Fig C); we obtained reportable data on about 3000 institutions. Overall, 56% (n=1,679) of institution responses indicated noncompliance with the HIPAA right of access, with 783 of those institution responses (47%) indicating noncompliance in two or more categories. Similar to results we saw on our scorecard analysis when submitting actual patient requests, refusal to send records to patients electronically by email was a primary reason for likely noncompliance. In the survey, noncompliant fee responses was the second highest reason for potential noncompliance.

**Figure C:**
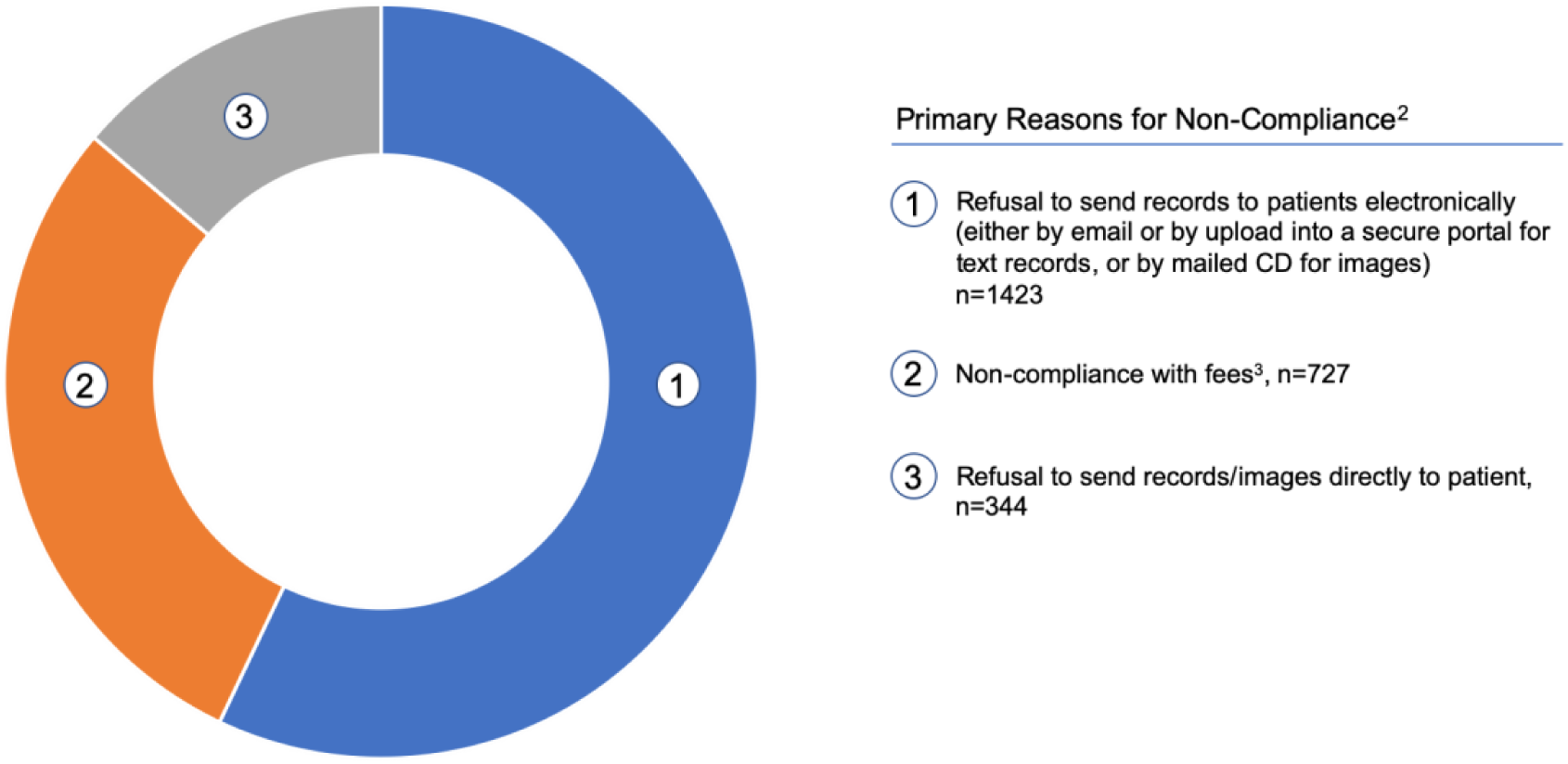
Likely Non-Compliant Institutions^1^ n=1,679 (56%)

### Notes

1. Non-compliance in at least one category indicates overall non-compliance. This is because all components are legally required to be compliant
2. Institutions can be non-compliant for more than one reason
3. We also found that of the 727 institutions non-compliant with fees, 521 (72%) were also non-compliant in another category

A number of institutions route patient access requests for images directly to their Radiology Departments or Radiology Film Libraries (collectively, “Radiology”). In comparing the responses of medical records departments to those of Radiology, medical records department responses were over four times more noncompliant than those of Radiology. However, of the 344 institutions whose responses indicated noncompliance regarding willingness to send information directly to the patient (responding they would send information only to another provider), 77% of the noncompliant responses came from Radiology.

Most institutions (n=2,616, 87%) responded that they would accept a request from patients sent by email or fax.

In 2016, OCR released extensive guidance on the HIPAA right of access that included a significant emphasis on fees. [14] We speculated that if an institution’s answer to the fee question indicated noncompliance, it was likely the institution was noncompliant in another category, using the fee issue as a proxy for whether the institution was generally up-to-date on their HIPAA right of access obligations. Our survey showed 72% (521/727) of providers whose responses indicated noncompliance with the fee provisions also had responses indicating noncompliance with another aspect of the right of access.

## Discussion

The privacy regulations under HIPAA have always included a right of individuals to access and receive copies of their complete medical records, with rare exceptions. In the Health Information Technology for Economic and Clinical Health Act of 2009 (HITECH), Congress clarified that individuals have the right to digital copies of electronic health records and to have those copies sent directly to a designated third party, such as a personal health record service or mobile health application (app). [17]] HHS incorporated the HITECH changes into the HIPAA Privacy Rule in 2013. [18] These changes to the HIPAA Privacy Rule right of access were part of an emphasis in HITECH on digital collection and exchange of health information and were expected to spark the development of more widespread personal health record services and mobile applications designed for use by individuals.

Notwithstanding the long history of this right, individuals have long struggled to exercise it. Inability to exercise the right of access has always been one of the top five categories of complaints to the OCR.[19] In 2016, complaints about inability to access records were, for the first time, the top category of complaints, surpassing inappropriate uses and disclosures for the first time.[19]

Customarily, individuals seeking copies of their medical records have had to submit requests on paper (or digitized paper) forms to medical records departments. The records, when produced, would be mailed (or sometimes faxed) to patients, or placed on a CD. Individuals often were required to pay fees – sometimes significant fees – to obtain their records. [20] However, recent federal efforts are pushing in the direction of enabling individuals to seamlessly access their health information online. Beginning with portals in certified electronic health record systems, coupled with incentive payments for providers to make data available to patients in those portals, and extending to more recent proposals for individuals to have an increasing amount of their health information available to them, via the app of their choice, through open standard application programming interfaces (APIs) and potential penalties for “blocking” information access by patients, the future of patient empowerment through seamless access to their health information is in sight.

These efforts – while promising – will take years to fully implement. The proposed timeline to implement APIs is two years after a final rule is published, and health care providers and EHR vendors are asking for more time.[21] Today, portals in EHRs are required to expose the data comprising the Common Clinical Dataset. [22] This is a good set of data – but it is significantly shy of all of the information that an individual has a right to under HIPAA. For example, it does not include images, notes, pathology reports, genomic/genetic test data. Federal officials have announced a glide path for expanding the data required to be accessible to patients via APIs - but this process will take years.[23] Consequently, patients seeking copies of all of their health records likely will need to obtain those records through a combination of digital access through APIs and the traditional route of submitting requests to medical records departments, which makes compliance with the HIPAA right of access by medical records departments and Radiology of continuing importance.

The scorecard and survey data collectively demonstrate that we have a long way to go to achieve consistent, seamless and HIPAA compliant processes for getting records to patients. As seen below, the results from the broad survey of nearly 3000 institutions, and the actual responses to patient requests by 210 providers, yielded similar results:

|  | Scorecard | Survey |
| --- | --- | --- |
| Overall Noncompliance/Indicated Noncompliance | 51% | 56% |
| % of Noncompliant (or Likely Noncompliant) Providers Refusing to Send Records Electronically by Email | 86% | 85% |

One distinction between the survey and scorecard results worth noting: the institutions’ responses on the survey indicated that 24 percent were likely noncompliant with the fee provisions of the HIPAA Right of Access. However, only two of the 210 providers evaluated in the scorecard were noncompliant due to noncompliant fees. Ciitizen spends time on the phone with medical records staff, supervisors and privacy officials at scorecard providers (and their records vendors) to ensure the requests are processed in compliance with HIPAA, which could explain why only lawful fees have been charged.

It appears from this study that training of medical record department and Radiology staff is critical to ensuring that patient requests are processed consistently in accordance with the HIPAA Privacy Rule. That more than half of providers are noncompliant – and that it takes multiple phone calls with supervisors or privacy officials for some providers to be compliant - strongly suggests that the average patient - not necessarily armed with textbook knowledge of the HIPAA requirements - would likely be far less successful at getting their requests processed in compliance with the law and might give up due to lack of time or frustration. (OCR Director Roger Severino publicly shared that he gave up on his efforts to obtain his own medical records. [24]) In particular, the requirement to send information to patients (or their designees) in the form or format the patient requests – including by email (or CD for images) – is an aspect of the HIPAA right of access that needs to be reinforced. Although overall the actual performance by scorecard providers on the fee limitations was much better than expected given the survey data, this is still an area that providers also need to evaluate to assure they are in compliance with the law.

In conclusion, with more than 50% of providers either out of compliance or at significant risk of noncompliance, the rights of patients to their health records is still being violated by too many health systems. Although many entities, including ONC and OCR, are working to educate patients and providers, additional enforcement of the right of access by OCR is needed.

We engaged in this study not to name and shame but to educate hospitals and other providers on the extent of noncompliance with the HIPAA Right of Access that exists – and the need for all HIPAA covered entities to examine their processes and ensure compliance with the HIPAA Right of Access.

We also wish to highlight for policymakers just how difficult it continues to be for patients to access their health information. Efforts to digitize this process have been proposed, but it will be years before seamless digital access by patients to all of their health information is a reality. In the meantime, requests to medical records departments (and Radiology) will still be required to enable patients to amass all of their health information. It is critical that these processes be compliant with HIPAA and responsive to patient needs.

## Ethical Review

The scorecard and survey do not constitute human subjects research under HIPAA or the Common Rule. The scorecard is retrospectively evaluating the responses of health care institutions to Ciitizen users’ requests for medical records that were processed by Ciitizen staff. Ciitizen users expressly consented to Ciitizen assisting in the gathering of their medical records; also, as part of the consent that each user executes to open a Ciitizen account, users were required to assent to Ciitizen’s privacy policy, which makes clear that Ciitizen can publish aggregate statistics about use of Ciitizen services.[25] The survey retrospectively evaluated institutional policies on patient record access based on telephone responses; the survey was evaluating the institutions, not the individuals responding to the call.

## Limitations

This study has several limitations. The survey data was gathered by Ciitizen staff, including temporary staff added just for purposes of compiling the survey data. We instructed all surveyors to use the script (see Box 2) but we did not record the calls, nor were the surveyors supervised or monitored in making these calls. Also, because the responses on fees were so varied, we did not have conventions/standards for surveyors to follow in recording their information. We also acknowledge that responses could be mis-recorded by staff. These are all reasons why we are reporting the survey data as *indicating* compliance or noncompliance. Given that the information we received in the survey could have been provided to any patient randomly calling with the same questions, we still believe the survey results could be instructive to hospitals in terms of ensuring that proper information about HIPAA right of access processes is being provided to the public.

The scorecard was created for this study and is not an established instrument; it was based on actual patient requests to health care providers. Here our involvement to get some of these requests processed was extensive, as often the requests needed to be escalated. This suggests that the experience of a patient, without any help, could have been worse. Institutions and medical practices did not realize at the time that they were being evaluated on their processes, so it is appropriate to consider this as an experience that could have happened to any patient.

In both the scorecard and survey, we listed providers separately by location. Although health care providers have the option under the HIPAA Privacy Rule to consolidate HIPAA compliance responsibilities for all of their locations under a central office [26], they are not required to do so – and unraveling those corporate relationships, which often change, would have been difficult, if not impossible, to do. Since the experience of a patient in requesting their health information is to query the location where they received care, we believe the scorecard and survey more accurately represents what a patient would experience if they made an access request or inquired about making one.

## Data Availability

The url with supplemental data referred to in the manuscript will be available November 12, 2019.

https://www.ciitizen.com/scorecard

